# Immunogenetics of Gastrointestinal Cancers: A Systematic Review of Inborn Errors of Immunity in Humans

**DOI:** 10.1101/2022.01.25.22269823

**Authors:** Beishi Zheng, Howard Chung, Chen Bing, Siming Sun, Peter H.R. Green, Timothy C. Wang, Xiao-Fei Kong

## Abstract

**Background/Aims:** The immune system plays a key role in cancer surveillance and modulation of the tumor microenvironment. Humans with inborn errors of immunity (IEI), or primary immunodeficiencies, may be prone to recurrent mucosal bacterial and viral infections and chronic inflammation, associated with intrinsic or secondary epithelium dysfunction, a potential risk factor for early-onset gastrointestinal (GI) cancer.

**Methods:** We systematically reviewed all cases with clinical diagnoses of both an IEI and a GI cancer in three databases (MEDLINE, SCOPUS, EMBASE). In total, 76 publications satisfying our inclusion criteria were identified, and data for 149 cases were analyzed.

**Results:** Of the 149 patients with IEIs, 95 presented with gastric cancer, 13 with small bowel cancer, 35 with colorectal cancer, and six with unspecified cancer or cancer at another site. Gastric and colonic adenocarcinoma was the most common. For both gastric and colorectal cancer, age at onset was significantly earlier in patients with IEIs than in the general population, based on the SEER database. Common variable immune deficiency (CVID) was the most common IEI associated with gastrointestinal cancer. About 12% of patients had molecular genetic diagnoses, the three most frequently implicated genes being *ATM, CARMIL2, CTLA4*. Impaired humoral immunity and Epstein-Barr virus (EBV) infection were frequently reported as the factors potentially underlying early-onset GI malignancy in patients with IEIs.

**Conclusion:** Patients with IEIs should be considered at risk of early-onset GI cancers, and should therefore undergo cancer screening at an earlier age. Surveillance guidance based on stratifications for genetic risk should be revised to take into account the immunogenetic contribution to GI cancers.

## Introduction

Cancer immunosurveillance, first proposed by Frank Macfarlane Burnet and Lewis Thomas in 1957, plays an indispensable role in the prevention of gastrointestinal cancers ^1^. Cancers were found to occur more frequently in immunocompromised individuals in studies of AIDS patients or organ recipients, particularly for non-melanoma skin cancer ^2, 3^. Studies of transgenic mice with defects of crucial components of the immune system have improved our understanding of the effector molecules involved in immunosurveillance and led to the development of immune checkpoint inhibitors, a key breakthrough in immunotherapy for cancer treatment ^4, 5^. Inborn errors of immunity (IEIs), also known as primary immunodeficiencies, manifest as recurrent infections, autoimmunity, autoinflammatory diseases, allergy, and cancer ^6^. Humans with IEIs provide us with a unique angle of study, or an experiment in nature, for elucidating immunosurveillance for GI cancer and the interaction between environment and host. However, the rare cases of GI cancer in patients with IEIs are challenging for the physicians responsible for their care. For instance, IEIs can manifest as an early-onset GI cancer, but unlike high-risk hereditary risk factors for gastrointestinal cancer, IEIs are not yet generally considered as a risk factor for GI cancers, and there are currently no practice guidelines for screening for gastrointestinal cancers in patients with IEIs ^7, 8^.

The diagnosis of an early-onset (before the age of 50 years) gastrointestinal cancer, whether gastric or colorectal cancer, is devastating for any patient ^9-11^. The field of primary immunodeficiencies has progressed considerably with recent advances in genetic sequencing technology, which are facilitating genetic diagnosis and discoveries. More than 400 genes have been shown to cause a wide array of immunodysregulation, with a broad spectrum of clinical presentations, some dominant and others recessive, and complete or incomplete penetrance ^6^. Epidemiological data suggest that inborn errors of immunity are more prevalent than previously thought, affecting about 1:8,500 to 1:100,000 individuals in the general population ^12^. With earlier clinical recognition, appropriate treatments, and hematopoietic stem cell transplantation, many patients with IEIs now survive into adulthood, overcoming severe infections or inflammation. Given these improvements in the overall survival of patients with IEI, physicians may begin to see more delayed clinical phenotypes with an incomplete penetrance, including gastrointestinal cancers. It is often difficult to understand the human genetics and immunologic basis of IEIs, due to phenotypic and allelic heterogeneity, differences in expressivity and incomplete penetrance. Given the complexity of human genetics and immunology, and the rareness of individuals with identified IEIs and gastrointestinal cancer, we systematically reviewed all the reported cases with both an IEI and a gastrointestinal cancer. We summarized their genetic and immunological features and highlighted the elements of importance for the clinical surveillance and management of this special population.

## Methods

### Case identification and literature search

We followed a Preferred Reporting Items for Systemic Reviews and Meta-Analysis (PRISMA)^13^ flow chart for the literature search and data collection for this systematic review (Supplementary Figure 1). Briefly, we conducted a search of the MEDLINE, SCOPUS, EMBASE databases for all publications up to December 1, 2020, focusing on IEIs and GI cancers. We focused on luminal GI cancers, and excluded pancreatic or hepatobiliary cancers. We followed the International Union of Immunological Societies (IUIS) guidelines ^6^ to ensure the inclusion of all IEIs. We searched three databases for the following terms: “ gastrointestinal neoplasms”, “ primary immunodeficiency”, “ agammaglobulinemia”, “ common variable immunodeficiency”, “ dysgammaglobulinemia”, “ lymphopenia”, “ immune dysregulation” and 14 additional Medical Subject Headings (MeSH) terms relating to IEIs in the title, abstract, or keywords of articles (Supplementary Table 1). The results of three database searches (*n*=1146) were then carefully reviewed, as illustrated in Supplementary Figure 1. Additional publications (*n*=24) were identified and included manually if missed by the search strategy described above search. Duplicated articles were consolidated. All records (*n*=1099) were then reviewed independently by two authors (B.Z and H.C) by comprehensive reading, and any discrepancies were resolved by a third author (X-F.K). The inclusion criteria were as follows: 1) Clinical diagnosis of IEI for cases; 2) Clinical diagnosis of gastrointestinal cancers for cases. Exclusion criteria were: 1) Not written in English; 2). Lack of detailed clinical information, such as age, sex, and age-at-onset data for the patient. In total, 76 articles satisfied with our inclusion criteria and had no exclusion criteria.

### Data curation, quality assessment and statistical analysis

We collected information by scrutinizing the publications identified to generate a dataset with the following information: author and year of publication of the article and, for the patients, sex, ethnicity, age at cancer diagnosis, genetic defects, immunological dysfunctions, IEI diagnosis, cancer histology and prognosis. Standardized forms were used to extract the required information from the articles. A quality assessment form was developed based on the CARE (CAse REports)^13^ tool for quality assessment (Supplementary Table 2). The expected distributions of age, sex, and survival interval of GI cancers were calculated from the data in the SEER (Surveillance, Epidemiology and End Results) database. Time-to-event and survival analyses (Log rank) were performed in GraphPad Prism 8.0 to calculate the hazard ratio. Categorical variables were analyzed with a Fisher’s exact test or *Chi*-squared test. Continuous variables were analyzed with *t-*tests. All data were analyzed with the statistical packages in R software (4.0.0).

## Results

### 1. A clinical summary of GI cancers in patients with IEIs

The 76 publications identified ^14-89^ included 149 cases in which both IEIs and gastrointestinal cancers have been diagnosed. A summary of the clinical characteristics of the patients concerned is provided in Figure 1. Briefly, 95 cases were diagnosed with gastric cancer, 13 cases presented with small bowel cancer, 35 cases with colorectal cancer, three cases with esophageal cancer, and three cases with a GI tract tumor unspecified location (Figure 1A). In total, 59 cases (40%) were reported after 2010 (Figure 1B). The dataset consisted of 84 (56.4%) male patients, 58(38.9%) female patients and seven patients for whom sex was not specified (Figure 1C). Age at onset for the cancer ranged from three to 82 years, with a median age at onset of 42 years (Figure 1D). Most of the reported cases (64.4%) were diagnosed before the age of 50 years and their cancers should therefore be considered early-onset. Age at onset for GI was normally distributed, contrasting with the left-skewed distribution in the general population. Fisher’s exact test showed that the proportion of male patients was higher in the younger age groups (ages 0-40) than the older age groups (ages above 40, p=0.034) (Figure 1D). Age at onset of the GI cancer was 26 to 30 years lower for the individuals with IEIs than for the general population (42 vs 68-72 years), according to the data of the SEER database. Ethnicity data were available for only 18 patients: seven white, four non-Hispanic, three Asian, two African American, and two Hispanic or Latino individuals. The IEI diagnosis was common variable immunodeficiency (CVID) for 88 patients in the cohort. The other IEI diagnoses included X-linked agammaglobulinemia (XLA) in 11 patients, selective IgA deficiency (SIgAD) in 10 patients, and ataxia-telangiectasia mutated gene deficiency (*ATM*) in 10 patients. The rate of molecular genetic diagnosis for IEIs was low, with disease-causing genotypes reported for only 18/149 (12.1%).

**Figure 1:**
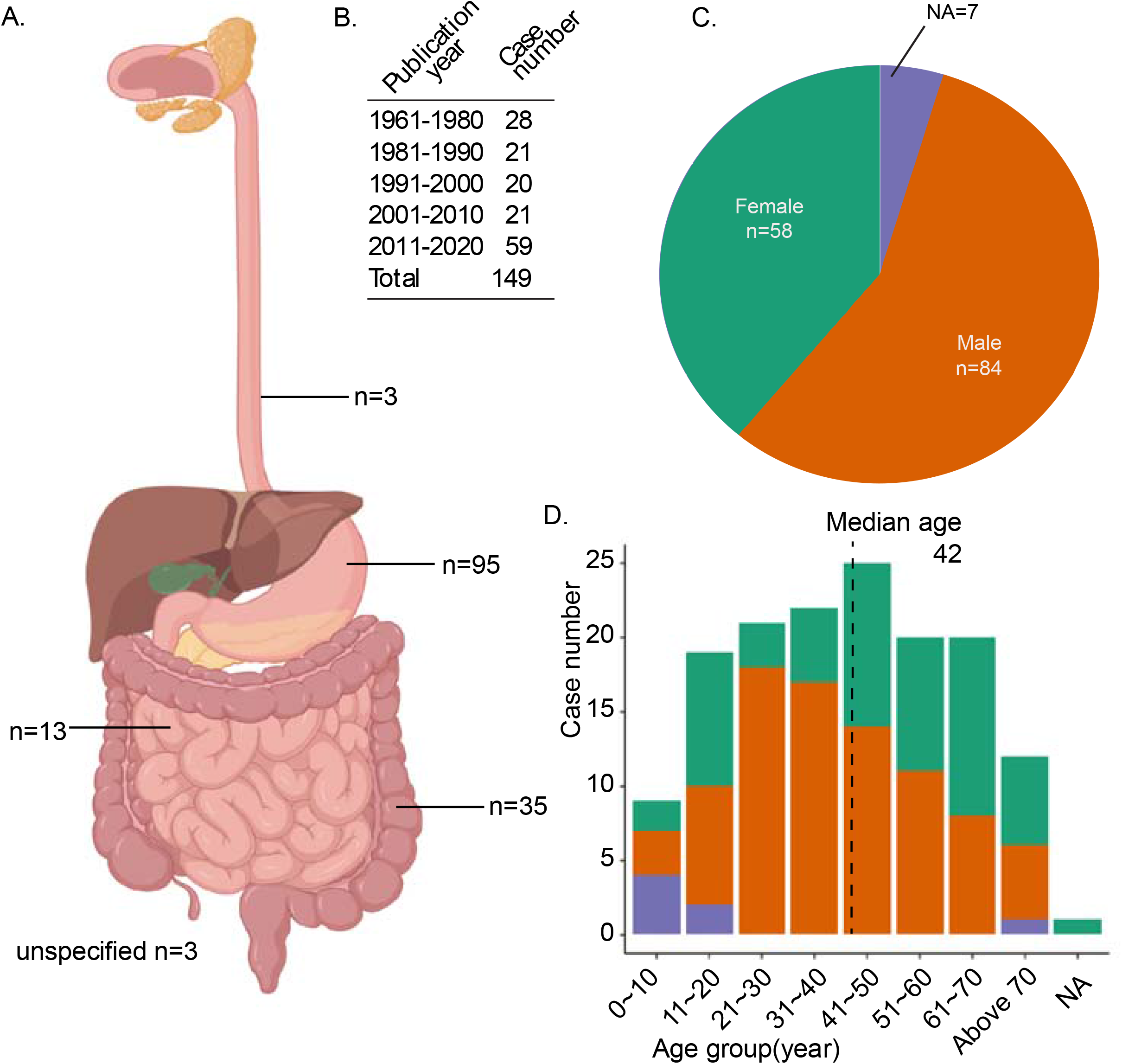
A descriptive summary of patients with inborn errors of immunity and GI cancers. A. A schematic presentation of the numbers of esophageal, gastric, small bowel and colon cancers reported in patients with IEIs. B. The table describes the number of cases published over the last six decades. C. A pie chart of the sex distribution for this cohort. D. A histogram presents the age and sex distributions of the cohort. NA: Not available.

### 2. Gastric cancers in patients with IEIs

Our literature search identified 95 patients with IEIs and gastric cancer, which was the most common GI cancer (Supplementary Table 3). Most (87/95, 91.5%) of the patients were diagnosed with gastric adenocarcinoma. The other cancer diagnosed were lymphoma (7/95, 7.4%), and neuroendocrine carcinoma (1/95, 1.1%) (Figure 2A). The mean age at gastric cancer diagnosis was 45.86 years, male patients outnumbered female patients in this group (55.8% vs 43.2%), and there were no significant differences with respect to the expected value (Figure 2B). Age at onset of gastric cancer in patients with IEIs was significantly different from that in the general population, with most patients with IEIs diagnosed with gastric cancer before the age of 65 years (Figure 2B). Gastric cancer was diagnosed between the ages of 25 and 44 years in 32.6% of IEI cases, whereas such early diagnosis was uncommon in the general population (Figure 2B). A time-to-event data analysis based on the observed and expected age at onset for gastric cancer yielded a hazard ratio (HR) of 3.57 for the IEI group, indicating a significantly earlier than expected age at onset for this group (Figure 2C). Prognosis was as follows: 31 (32.6%) patients died within one year, and nine (9.5%) patients died within five years of a gastric cancer diagnosis. The five-year survival rate in IEI patients with gastric cancer was 27.3%, which is close to the value reported for the general population, as shown by the survival analysis (Figure 2D). CVID was the most common IEI and was found in 69 cases (72.6%) of gastric cancer. *ATM* defects and XLA were the next most frequent IEIs (Figure 2E). Four patients with *CTLA4* deficiency and one patient with *LRBA* deficiency were reported to have gastric cancer. However, most of the publications did not report the history of *Helicobacter pylori* infection. In summary, inborn errors resulting in B-cell defects, or dysfunctions of humoral immunity were the most common IEIs associated with gastric cancer.

**Figure 2:**
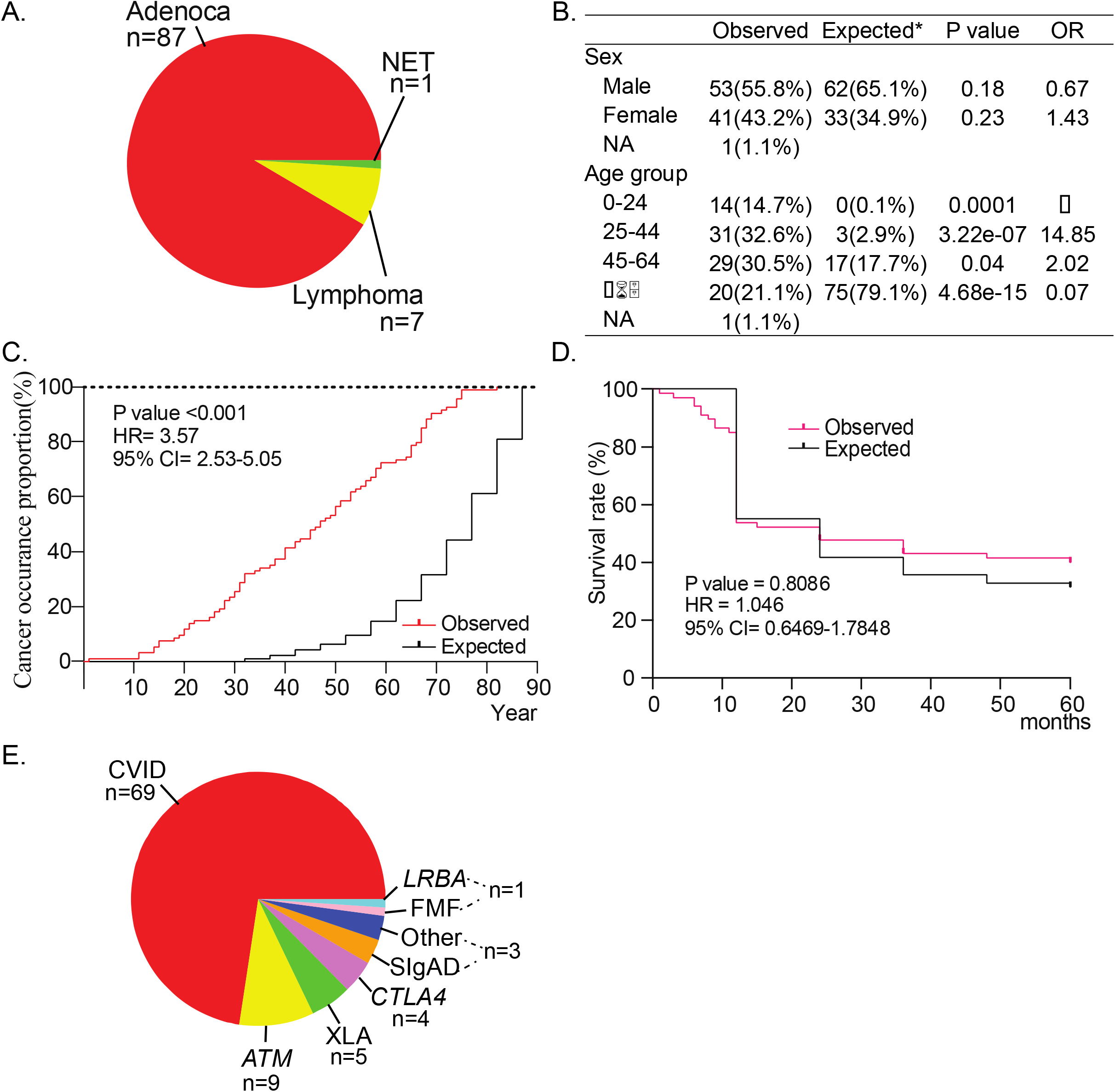
Gastric cancers reported in patients with IEIs. A. Cellular types of gastric cancer reported in patients with IEIs. Adenoca: Adenocarcinoma, NET: Neuroendocrine tumor. B. The table shows the sex and age distributions of gastric cancer patients from the IEI cohort relative to the expected numbers in the general population imputed from the SEER database. OR: Odds ratio. C. A pie chart of the different types of IEI and numbers of cases with gastric cancer. CVID: Common variable immune deficiency; ATM: Ataxia-telangiectasia mutated, XLA: X-linked agammaglobulinemia, CTLA4: Cytotoxic T-lymphocyte associated Protein 4, SIgAD: Selective IgA deficiency, FMF: Familial Mediterranean fever, LRBA: LPS responsive beige-like anchor protein. D. Survival curve for gastric cancer comparing patients with and without IEIs. NA: Not available.

### 3. Small bowel cancers reported in patients with IEIs

The small bowel is an uncommon site for cancers. To date, 13 patients with IEIs and small bowel cancer have been reported (Supplementary Table 4). Lymphoma was the most common type of cancer (11/13, 84.6%) affecting the small bowel, with 10 cases of B-cell lymphoma, one of T-cell lymphoma. Neuroendocrine carcinoma and reticular lymphosarcoma were reported in one case. Mean age at cancer diagnosis was 43.92 years and there were seven male patients and six female patients. CVID (41.6%, 6/13) was the most common IEIs associated with small bowel cancer, consistent with the recent meta-analysis of CVID patients showing that lymphoma was the cancer most frequently reported in these patients ^90^. One patient with Wiskott-Aldrich syndrome and another with familial Mediterranean fever were reported. Overall, 41.6% (6/13) of the patients survived for more than five years, 15.4% (2/13) survived for less than three years, and 23.1% (3/13) died within one year. The genetic etiology of small-bowel lymphoma in CVID remains to be explored.

### 4. Colorectal cancers in patients with IEIs

There were 35 cases of colorectal cancer reported in patients with IEIs (Supplementary Table 5). The most common type of cancer was adenocarcinoma (68.1%, 24/35), followed by lymphoma (11.4%, 4/35), and smooth-muscle tumors (5.7%, 2/35) (Figure 3A). Other cellular types of cancer were reported in one case each, including neuroendocrine carcinoma, rhabdomyosarcoma, and squamous cell carcinoma (Figure 3A). Mean age at onset was 32.94 years, and this group comprised 60% (21/35) male and 28.6% (10/35) female patients. Most of the cases (71.4%, 25/35) were diagnosed before 45 years of age, which is significantly younger than expected (Figure 3B). Time-to-event analysis (Figure 3C) revealed a HR of 5.23 (95% CI 2.65-10.30) for the IEI group. Overall, 42.9% (15/35) of the patients with IEIs died within one year of colon cancer diagnosis, and five-year survival rate for IEIs patients with colon cancer was 31.4% (11/35). A survival analysis showed prognosis to be worse for the IEI group than for the general population (HR = 2.55, *p* = 0.002) (Figure 3D). The most common immune dysregulations were B-cell defects (71.4%, 25/35), including CVID, XLA or selective IgA deficiency. However, DNA repair defects, including *BLM, NBS1* and *ATM* gene mutations, accounted for 20% (7/35) of IEI cases with colorectal cancer (Figure 3E). In summary, colorectal cancer in patients with IEIs appeared to be more heterogeneous in terms of immunological dysfunction and to lead to worse outcomes than such cancers in the general population.

**Figure 3:**
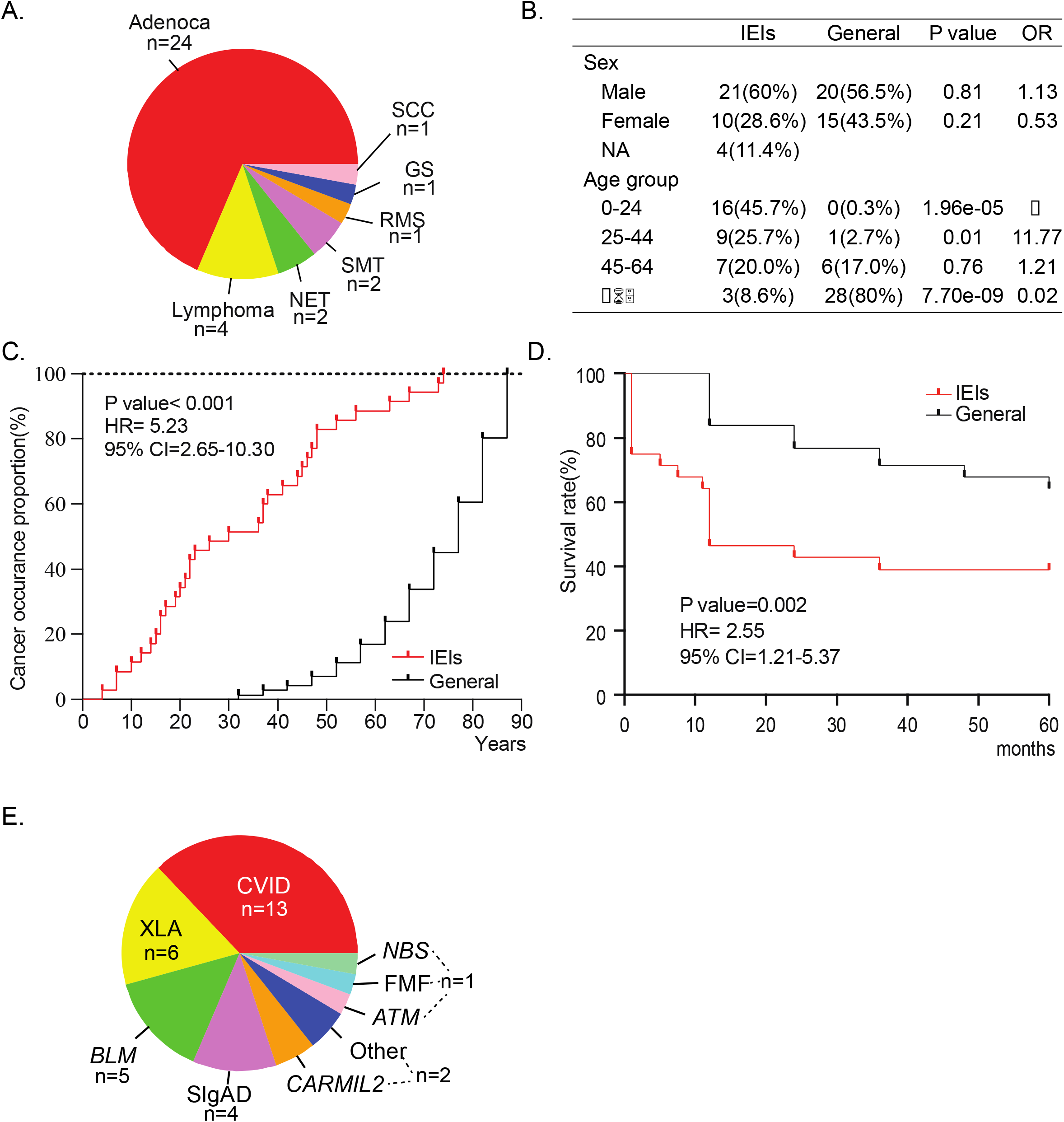
Colon cancer in patients with IEIs. A. Cellular types of colon cancer reported in patients with IEIs. Adenoca: Adenocarcinoma, NET: Neuroendocrine tumor, SMT: Smooth muscle tumor, RMS: Rhabdomycosarcoma, GS: Granulocytic sarcoma, SCC: Squamous cell carcinoma. B. The table shows the sex and age distribution of colon cancer patients from the IEI cohort relative to the expected numbers in the general population imputed from the SEER database. OR: Odds ratio. C. A pie chart of the different types of IEIs and numbers of cases with colon cancer. CVID: Common variable immune deficiency; ATM: Ataxia-telangiectasia mutated, XLA: X-linked agammaglobulinemia, BLM: Bloom syndrome, SIgAD: Selective IgA deficiency, *CARMIL2*: Capping Protein Regulator And Myosin 1 Linker 2, ATM: Ataxia-telangiectasia mutated, FMF: Familial Mediterranean fever, NBS: Nijmegen breakage syndrome. D. Survival curve for colon cancer comparing patients with and without IEIs. NA: Not available.

## Discussion

The Fearon-Volgelstein model has prevailed over the last few decades, and genetic practices for GI cancer have now emerged and can facilitate the management of about 3% to 5% of all colorectal cancer patients^91^. Pathogenic variants with various degrees of clinical penetrance have been identified in 9-26% of patients with early-onset colorectal cancer ^92^. However, the detection of such mutations is limited by a lack of knowledge of the full genetic landscape for GI cancers. We investigated the immunogenetic determinants of GI cancers by performing a systemic review of cases in patients with IEIs, who usually have a genetic predisposition for cancer and abnormal immunological dysregulation. Even though IEIs are usually diagnosed during childhood and mortality rates are high at a young age, 2-10% of patients have been found to develop cancers with their short lifespans ^78^. Our analysis indicates that patients with IEIs may be prone to early-onset GI cancers. Longitudinal data from a large IEI cohort are lacking, but our findings are consistent with previous CVID studies ^58, 93^. Further studies are required to delineate the clinical epidemiology of IEIs in patients with early-onset GI cancers, through immunological or genetic testing. A collaboration between clinical immunologists and pediatricians might facilitate GI cancer surveillance in individuals with IEIs. We need to address the question as to whether there is an association between clinical symptoms or biomarkers of IEIs and early-onset GI cancer? It would be intriguing to determine whether immunoglobulin level, lymphocytopenia, a clinical diagnosis of CVID, or a history of IVIG use is associated with early-onset GI cancer, ideally based on data from a large registry of GI cancer patients.

Several factors could, conceivably, account for the early-onset gastrointestinal cancers observed in patients with IEIs. First, primary immunodeficiency may lead to chronic mucocutaneous infection (with *Helicobacter. pylori* or EBV, for example,) in the gastrointestinal tract, which is a risk factor for gastric cancer. *CARMIL2* deficiency has been associated with EBV^+^ smooth muscle tumors in the GI tract ^65^. Previous studies on CVID have revealed a high prevalence of *H. pylori* infection and multifocal atrophic gastritis, but *H*.*pylori* infection rates in CVID patients vary between reports and may not, necessarily, explain the risk of GI cancer ^67^. Nevertheless, one recent large CVID cohort meta-analysis ^90^ on 8123 CVID patients revealed an estimated prevalence of gastric cancer in CVID patients of about 1.5%, lower than the prevalence of 4.1% for lymphoma, consistent with another report suggesting that gastric cancer may be the leading cause of death in CVID patients ^67^. Gastrointestinal cancers may also arise due to defects of immune cells in the mucosa. As most reported cases have impaired humoral immunity, this suggests that B cells may play an important role in gastrointestinal mucosal surveillance. Only a few cases with genetic abnormalities have been reported, but several genetic disorders have well-defined B-cell abnormalities, including *CTLA4, LBRA*, and *CARMIL2* mutations.

A third plausible explanation is related to intrinsic epithelium abnormalities due to the pleiotropic effects of some genes. Three disorders, ataxia-telangiectasia due to *ATM* mutations, Nijmegen breakage syndrome due to *NBS1* defects, and Bloom syndrome due to *BLM* mutations, have been reported to cause heterogeneous clinical phenotypes and chromosomal instability. ATM is a serine/threonine kinase activated by DNA damage, it phosphorylates several tumor suppressors, including p53, NBS1, and H2AX. Eight patients (9.5%) with ataxia-telangiectasia due to biallelic *ATM* gene mutations have been reported to have gastric cancer and immunodeficiency ^33, 50, 76, 82, 83, 85^. Individuals with heterozygous germline *ATM* mutations might have an increased risk for early-onset gastric and pancreatic cancer ^94^. However, some IEIs characterized by impaired DNA, such as MCM4 or GINS1 deficiencies, have not been found to lead to cancers ^95, 96^. It is possible that GI cancers in patients with DNA repair disorders are more strongly related to intrinsic epithelial cell defects than to compromised immunity. With advances in human genetics and immunological technologies, studies of humans with IEIs might improve our understanding of immune surveillance in the gastrointestinal tract in general.

## Data Availability

All data produced in the present study are available upon reasonable request to the authors

## Acknowledgments

We thank Dr. Jean-Laurent Casanova, Laurent Abel, Anil K Rustgi and Benjamin Lebwohl for their careful reading of the manuscript and suggestions. Figure 1 was created with BioRender.com.

