## Supplementary material for "Immunogenetics of Gastrointestinal Cancers: A Systematic Review of Inborn Errors of Immunity in Humans"

**Supplemental file**

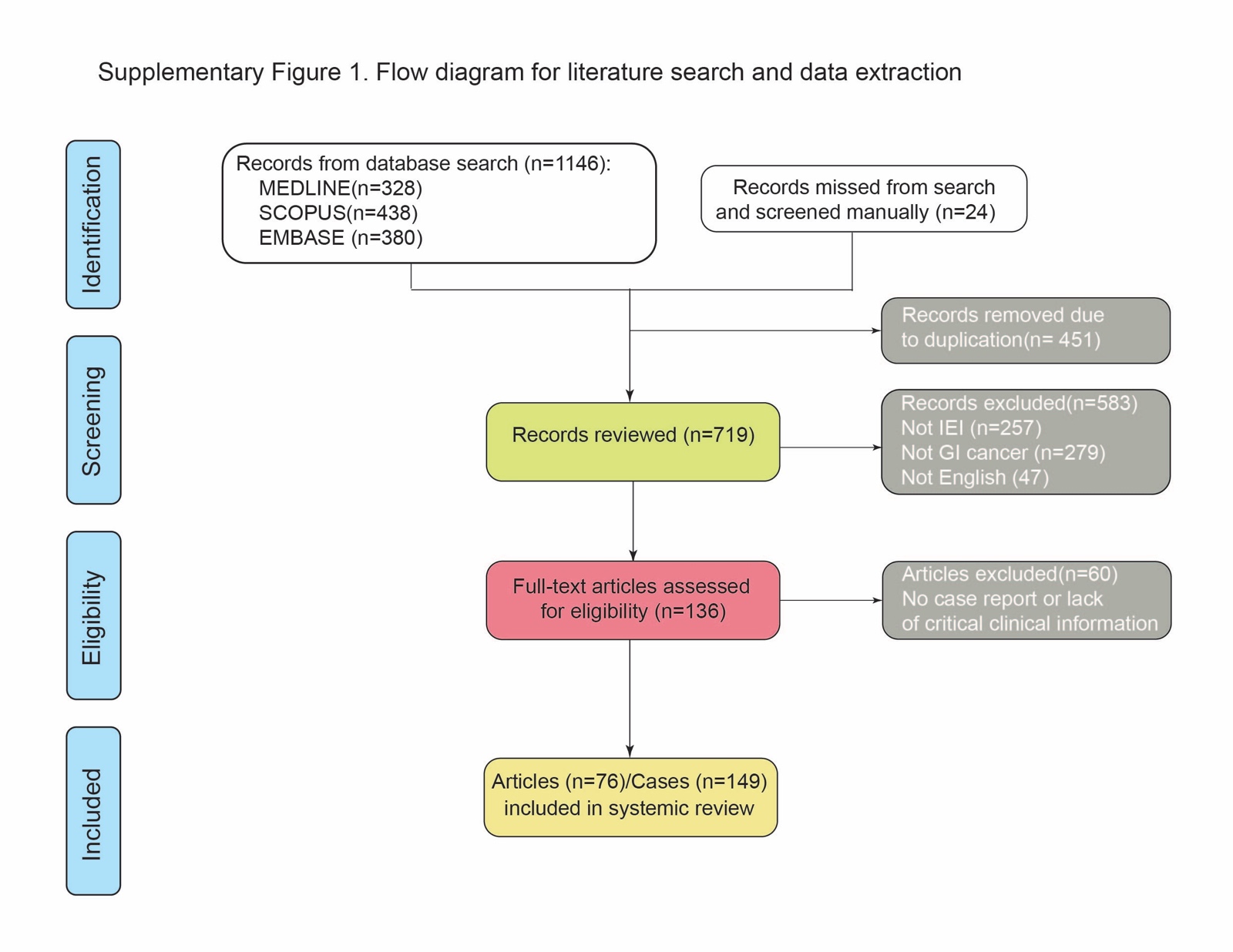

**Supplementary Table 1. Search Terms**

| Database | Search terms |
| --- | --- |
| MEDLINE | ((((((((("Agammaglobulinemia"[Mesh]) OR "Common Variable Immunodeficiency"[Mesh]) OR "Dysgammaglobulinemia"[Mesh]) OR "Lymphopenia"[Mesh]) OR "Phagocyte Bactericidal Dysfunction"[Mesh]) OR "Primary Immunodeficiency Diseases"[Mesh]) OR "Hereditary Autoinflammatory Diseases"[Mesh]) OR ("Gastrointestinal Neoplasms"[Mesh] AND "immune dysregulation"[TIAB])) OR (("Immunity, Innate"[Mesh]) AND "Immunologic Deficiency Syndromes"[Mesh]) AND "Gastrointestinal Neoplasms"[Mesh] |
| SCOPUS | ( title-abs-key ( "gastrointestinal neoplasms" )  AND  title-abs-key ( "primary immunodeficiency" ) ); ( title-abs-key ( "gastrointestinal neoplasms" )  and  title-abs-key ( "agammaglobulinemia" ) ); ( title-abs-key ( "gastrointestinal neoplasms" )  AND  title-abs-key ( "CVID" ) ); ( title-abs-key ( "gastrointestinal neoplasms" )  AND  title-abs-key ( "common variable immunodeficiency" ) ); ( title-abs-key ( "gastrointestinal neoplasms" )  AND  title-abs-key ( "dysgammaglobulinemia" ) ); ( title-abs-key ( "gastrointestinal neoplasms" )  AND  title-abs-key ( "lymphopenia" ) );( title-abs-key ( "gastrointestinal neoplasms" )  AND  title-abs-key ( "primary immunodeficiency diseases" ) ); ( title-abs-key ( "gastrointestinal neoplasms" )  AND  title-abs-key ( "immune dysregulation" ) ); ( title-abs-key ( "gastrointestinal neoplasms" )  AND  title-abs-key ( "immunologic deficiency syndromes" ) );( title-abs-key ( "stomach cancer" )  AND  title-abs-key ( "ATM" ) ); ( title-abs-key ( "colon cancer" )  AND  title-abs-key ( "ATM" ) ); ( title-abs-key ( "colon cancer" )  AND  title-abs-key ( "CVID" ) ); ( title-abs-key ( "stomach cancer" )  and  title-abs-key ( "CVID" ) ) |
| EMBASE | "gastrointestinal malignancy" AND ("primary immunodeficiency" OR "ATM" OR "Agammaglobulinemia" OR "Common Variable Immunodeficiency" OR "Dysgammaglobulinemia" OR "Lymphopenia" OR "Phagocyte Bactericidal Dysfunction" OR "Hereditary Autoinflammatory Diseases" OR "immune dysregulation") |

**Supplementary Table 2. Quality Assessment Form**

| Paper # | Reference first author | Year | IEI and GI cancer included in abstract | Tumor pathology provided | IEI type provided | Gene defect  provided | Prognosis provided | Clinical details provided |
| --- | --- | --- | --- | --- | --- | --- | --- | --- |
| 1 | Huizenga KA | 1961 | Yes | Yes | Yes | No | No | No |
| 2 | Kildeberg P | 1961 | Yes | Yes | Yes | No | No | No |
| 3 | Hermans PE* | 1966 | Yes | Yes | Yes | No | No | No |
| 4 | Miller DG | 1967 | Yes | Yes | Yes | No | No | No |
| 5 | Radl J | 1967 | Yes | Yes | Yes | No | No | No |
| 6 | Haerer A | 1969 | Yes | Yes | Yes | No | Yes | Yes |
| 7 | Fraser KJ | 1970 | Yes | Yes | Yes | No | Yes | Yes |
| 8 | Kersey JH* | 1973 | Yes | Yes | Yes | No | No | No |
| 9 | Hamoudi AB | 1974 | Yes | Yes | Yes | No | Yes | Yes |
| 10 | Faraci RP | 1975 | Yes | Yes | Yes | No | Yes | Yes |
| 11 | Hermans PE* | 1976 | Yes | Yes | Yes | No | Yes | No |
| 12 | Siegel SE | 1976 | Yes | Yes | Yes | No | Yes | Yes |
| 13 | Watanabe A | 1977 | Yes | Yes | Yes | No | Yes | Yes |
| 14 | Battle WM | 1978 | Yes | Yes | Yes | No | Yes | Yes |
| 15 | Frais M | 1979 | Yes | Yes | Yes | No | Yes | Yes |
| 16 | Lamers CB | 1980 | Yes | Yes | Yes | No | Yes | Yes |
| 17 | Gonzalez-Vitale JC | 1982 | Yes | Yes | Yes | No | Yes | Yes |
| 18 | Mir-Madjlessi SH | 1984 | Yes | Yes | Yes | No | Yes | Yes |
| 19 | Kinlen LJ* | 1985 | Yes | Yes | Yes | No | Yes | No |
| 20 | Aguilar FP | 1987 | Yes | Yes | Yes | No | No | Yes |
| 21 | Cunningham-Rundles C* | 1987 | Yes | Yes | Yes | No | Yes | No |
| 22 | Durham JC | 1987 | Yes | Yes | Yes | No | Yes | Yes |
| 23 | Takemiya | 1987 | Yes | Yes | Yes | No | Yes | Yes |
| 24 | Conley ME | 1988 | Yes | Yes | Yes | No | Yes | Yes |
| 25 | Cunningham-Rundles C | 1989 | Yes | Yes | Yes | No | Yes | No |
| 26 | Vorechovsky I | 1990 | Yes | Yes | Yes | No | Yes | Yes |
| 27 | Paraf F | 1991 | Yes | Yes | Yes | No | Yes | Yes |
| 28 | Adachi Y | 1992 | Yes | Yes | Yes | No | Yes | Yes |
| 29 | Castellano G | 1992 | Yes | Yes | Yes | No | Yes | Yes |
| 30 | Matz J | 1992 | Yes | Yes | Yes | No | Yes | Yes |
| 31 | de Bruin NC | 1993 | Yes | Yes | Yes | No | Yes | Yes |
| 32 | Lavilla P | 1993 | Yes | Yes | Yes | No | Yes | Yes |
| 33 | Ohnoshi T | 1993 | Yes | Yes | Yes | No | Yes | Yes |
| 34 | Van der Meer JM | 1993 | Yes | Yes | Yes | No | Yes | Yes |
| 35 | Cox JE | 1993 | Yes | Yes | Yes | No | No | Yes |
| 36 | Chiaramonte C | 1994 | Yes | Yes | Yes | No | No | Yes |
| 37 | Zirkin HJ | 1996 | Yes | Yes | Yes | No | Yes | Yes |
| 38 | Balci S | 1999 | Yes | Yes | Yes | No | Yes | Yes |
| 39 | Murphy RC* | 1999 | Yes | Yes | Yes | No | Yes | Yes |
| 40 | Wang J | 1999 | Yes | Yes | Yes | No | Yes | Yes |
| 41 | Zullo A | 1999 | Yes | Yes | Yes | No | No | No |
| 42 | Bachmeyer C | 2000 | Yes | Yes | Yes | No | Yes | Yes |
| 43 | Mellemkjael L | 2002 | Yes | Yes | Yes | No | No | No |
| 44 | Meyer S | 2004 | Yes | Yes | Yes | Yes | Yes | Yes |
| 45 | Lackmann GM | 2005 | Yes | Yes | Yes | No | Yes | Yes |
| 46 | Delia M | 2006 | Yes | Yes | Yes | No | Yes | Yes |
| 47 | Desar IME | 2006 | Yes | Yes | Yes | No | Yes | Yes |
| 48 | Gabizon I | 2006 | Yes | Yes | Yes | Yes | Yes | Yes |
| 49 | Daniels JA | 2007 | Yes | Yes | Yes | No | No | No |
| 50 | Dunnigan | 2007 | Yes | Yes | Yes | No | Yes | Yes |
| 51 | Purim O | 2007 | Yes | Yes | Yes | No | Yes | Yes |
| 52 | Brosens LAA | 2008 | Yes | Yes | Yes | Yes | No | Yes |
| 53 | Rosa DD | 2008 | Yes | Yes | Yes | Yes | No | Yes |
| 54 | Thomas ERA | 2008 | Yes | Yes | Yes | No | Yes | Yes |
| 55 | Malhotra | 2008 | Yes | Yes | Yes | No | No | Yes |
| 56 | Otabor IA | 2009 | Yes | Yes | Yes | No | Yes | Yes |
| 57 | Yap YL | 2009 | Yes | Yes | Yes | No | Yes | Yes |
| 58 | Benjazia E | 2011 | Yes | Yes | Yes | No | Yes | Yes |
| 59 | Dhalla F | 2011 | Yes | Yes | Yes | No | Yes | Yes |
| 60 | Slotta JE | 2011 | Yes | Yes | Yes | No | Yes | Yes |
| 61 | Turkbeyler IH | 2011 | Yes | Yes | Yes | No | Yes | Yes |
| 62 | Watkins C | 2012 | Yes | Yes | Yes | No | Yes | Yes |
| 63 | Patiroglu T | 2013 | Yes | Yes | Yes | No | Yes | Yes |
| 64 | Loh KP | 2014 | Yes | Yes | Yes | No | No | Yes |
| 65 | Staines Boone AT | 2014 | Yes | Yes | Yes | No | Yes | Yes |
| 66 | De Petris G | 2014 | Yes | Yes | Yes | No | Yes | Yes |
| 67 | Vlkova M | 2015 | Yes | Yes | Yes | No | No | No |
| 68 | Hayakawa S | 2016 | Yes | Yes | Yes | Yes | Yes | Yes |
| 69 | Goldberg A | 2016 | Yes | Yes | Yes | No | Yes | Yes |
| 70 | Bratanic N | 2017 | Yes | Yes | Yes | Yes | Yes | Yes |
| 71 | Schober T* | 2017 | Yes | Yes | Yes | Yes | Yes | Yes |
| 72 | Egg D | 2018 | Yes | Yes | Yes | Yes | Yes | Yes |
| 73 | Varricchi G | 2018 | Yes | Yes | Yes | No | Yes | Yes |
| 74 | Pulvirenti F* | 2018 | Yes | Yes | Yes | No | Yes | Yes |
| 75 | Antunes PDSL | 2019 | Yes | Yes | Yes | No | Yes | Yes |
| 76 | Li M | 2019 | Yes | Yes | Yes | Yes | Yes | Yes |

*Publications describe multiple cases with gastrointestinal cancers affecting different parts of the GI tract.

**Supplementary Table 3. Patients with gastric cancer**

| Paper # | Reference first author | Year | Type of cancer | Number of cases | Age at cancer diagnosis | Sex | IEI type | Gene defect | Immunodeficiency type | Prognosis |
| --- | --- | --- | --- | --- | --- | --- | --- | --- | --- | --- |
| 1 | Huizenga KA | 1961 | Gastric adenocarcinoma | 1 | 56 | 1F | CVID | NA | B-cell function deficiency | NA |
| 2 | Hermans PE | 1966 | Gastric adenocarcinoma | 1 | 31 | 1M | CVID | NA | B-cell function deficiency | NA |
| 3 | Haerer A | 1969 | Gastric adenocarcinoma | 2 | 20.5 | 2F | Ataxia telangiectasia | NA | Combined immunodeficiency | Pt1, 2 <1 year |
| 4 | Fraser KJ | 1970 | Gastric adenocarcinoma | 1 | 53 | 1M | IgA deficiency | NA | B-cell function deficiency | >5 years |
| 5 | Kersey JH | 1973 | Gastric adenocarcinoma | 4 | 59.25 | 2F, 2M | CVID | NA | B-cell function deficiency | NA |
| 6 | Hermans PE | 1976 | Gastric adenocarcinoma | 4 | 50 | 2F, 2M | CVID | NA | B-cell function deficiency | Pt1<3 years, Pt 2,3,4<1 year |
| 7 | Siegel SE | 1976 | Gastric adenocarcinoma | 1 | 1 | F | Ataxia telangiectasia | NA | Combined immunodeficiency | <1 year |
| 8 | Watanabe A | 1977 | Gastric adenocarcinoma | 1 | 14 | 1M | Ataxia telangiectasia | NA | Combined immunodeficiency | <1 year |
| 9 | Battle WM | 1978 | Gastric adenocarcinoma | 2 | 27.5 | 1F, 1M | CVID | NA | B-cell function deficiency | Pt1 <3 years, Pt2<1 year |
| 10 | Frais M | 1979 | Gastric adenocarcinoma | 1 | 26 | 1M | Ataxia telangiectasia | NA | Combined immunodeficiency | <1 year |
| 11 | Kinlen LJ | 1985 | Gastric adenocarcinoma | 8 | 53 | 3F, 5M | 7 CVID, 1 XLA | NA | B-cell function deficiency | 5 pt <1 year, 1 pt 5year, 3 pts >5 years |
| 12 | Cunningham-Rundles C | 1987 | Gastric adenocarcinoma | 1 | 66 | 1F | CVID | NA | B-cell function deficiency | >5 years* |
| 13 | Conley ME | 1988 | Gastric adenocarcinoma | 1 | 11 | 1F | CVID | NA | B-cell function deficiency | <1 year |
| 15 | Cunningham-Rundles C | 1989 | Gastric adenocarcinoma | 2 | 54 | 2F | CVID | NA | B-cell function deficiency | 2 NA |
| 15 | Vorechovsky I | 1990 | Gastric lymphoma | 1 | 32 | 1M | CVID | NA | B-cell function deficiency | <1 year |
| 16 | Paraf F | 1991 | Gastric lymphoma | 1 | 45 | 1F | Familial Mediterranean fever | NA | Autoinflammatory disorder | >3 years |
| 17 | Matz J | 1992 | Gastric adenocarcinoma | 1 | 37 | 1M | Common variable hypogammaglobulinemia | NA | B-cell function deficiency | <1 year |
| 18 | Lavilla P | 1993 | Gastric adenocarcinoma | 1 | 23 | 1M | XLA | NA | B-cell function deficiency | <5 years |
| 19 | Cox JE | 1994 | Gastric adenocarcinoma | 1 | 46 | 1M | CVID | NA | B-cell function deficiency | NA |
| 20 | Murphy RC | 1999 | Gastric lymphoma | 2 | 12.5 | 2F | Ataxia telangiectasia | NA | Combined immunodeficiency | NA |
| 21 | Zullo A | 1999 | Gastric adenocarcinoma | 1 | 65 | 1M | CVID | NA | B-cell function deficiency | NA |
| 22 | Bachmeyer C | 2000 | Gastric adenocarcinoma | 1 | 28 | 1M | XLA | NA | B-cell function deficiency | <1 year |
| 23 | Mellemkjaer L | 2002 | Gastric adenocarcinoma | 5 | 71.2 | 1F,4M | 2 IgA deficiency, 3 CVID | NA | B-cell function deficiency | NA |
| 24 | Lackmann GM | 2005 | Gastric adenocarcinoma | 1 | 15 | 1M | XLA | *BTK* | B-cell function deficiency | >5 years* |
| 25 | Delia M | 2006 | Gastric lymphoma | 1 | 28 | 1M | CVID | NA | B-cell function deficiency | >5 years* |
| 26 | Desar IME | 2006 | Gastric MALT | 1 | 65 | 1M | CVID | NA | B-cell function deficiency | >5 years* |
| 27 | Daniels JA | 2007 | Gastric adenocarcinoma | 1 | 67 | 1F | CVID | NA | B-cell function deficiency | NA |
| 28 | Malhotra RK | 2008 | Gastroenteropancreatic neuroendocrine carcinoma | 1 | 21 | 1M | X-linked hyperimmunoglobulin | NA | B-cell function deficiency | NA |
| 29 | Otabor IA | 2009 | Gastric adenocarcinoma | 1 | 20 | 1F | Ataxia telangiectasia | NA | Combined immunodeficiency | <1 year |
| 30 | Yap YL | 2009 | Gastric adenocarcinoma | 1 | 29 | 1M | CVID | NA | B-cell function deficiency | >5 years* |
| 31 | Slotta JE | 2011 | Gastric adenocarcinoma | 1 | 15 | 1M | Autosomal recessive agammaglobulinemia | μ heavy chain | B-cell function deficiency | >5 years* |
| 32 | Dhalla F | 2011 | Gastric adenocarcinoma | 1 | 75 | NA | CVID | NA | B-cell function deficiency | NA |
| 33 | Patiroglu T | 2013 | Gastric adenocarcinoma | 1 | 18 | 1F | Ataxia telangiectasia | NA | Combined immunodeficiency | NA |
| 34 | Loh KP | 2014 | Gastric adenocarcinoma | 1 | 58 | 1F | CVID | NA | B-cell function deficiency | NA |
| 35 | Staines Boone AT | 2014 | Gastric adenocarcinoma | 1 | 30 | 1M | XLA | *BTK* | B-cell function deficiency | <1 year |
| 36 | De Petris G | 2014 | Gastric adenocarcinoma | 6 | 46.3 | 4F, 2M | CVID | NA | B-cell function deficiency | 1 pt <5 years, 1 pt >5 years*,2 pt>5 years, 2 NA |
| 37 | Vlkova M | 2015 | Gastric adenocarcinoma | 1 | 75 | 1F | CVID | NA | B-cell function deficiency | NA |
| 38 | Hayakawa S | 2016 | Gastric adenocarcinoma | 1 | 35 | 1M | CTLA4 deficiency | *CTLA4* | T-cell function deficiency | <1 year |
| 39 | Bratanic N | 2017 | Gastric adenocarcinoma | 1 | 19 | 1M | LRBA deficiency | *LRBA* | T-cell function deficiency | NA |
| 40 | Egg D | 2018 | Gastric adenocarcinoma | 3 | 33.7 | 2M,1F | CTLA4 deficiency | *CTLA4* | T-cell function deficiency | 2 pt > 5 years, 1NA |
| 41 | Varricchi G | 2018 | Gastric adenocarcinoma | 1 | 58 | 1F | CVID | NA | B-cell function deficiency | NA |
| 42 | Pulvirenti F | 2018 | 25 Gastric adenocarcinomas, 1 MALT | 26 | 52.5 | 10F,16M | CVID | NA | B-cell function deficiency | 10 pt <1 year, 5 pt<5 years, 1 pt>5 years, 10 pt>5 years* |

*Patients still alive at the time of publication were considered to have a survival of more than 5 years

**Supplementary Table 4. Patients with small-bowel cancers**

| Paper# | Reference first author | Year | Type of cancer | Case number | Age at cancer diagnosis | Sex | IEI type | Gene defect | Immune deficiency type | Prognosis |
| --- | --- | --- | --- | --- | --- | --- | --- | --- | --- | --- |
| 1 | Radl J | 1967 | Reticular lymphosarcoma | 1 | 14 | 1M | Wiskott-Aldrich syndrome | NA | T-cell function deficiency | <3 years |
| 2 | Faraci RP | 1975 | Jejunal lymphoma | 1 | 8 | 1M | Wiskott-Aldrich syndrome | NA | T-cell function deficiency | <1 year |
| 3 | Lamers CB | 1980 | Jejunal lymphoma | 1 | 44 | 1F | Hypogammaglobulinemia | NA | B-cell function deficiency | >5 years* |
| 4 | Gonzalez- Vitale JC | 1982 | Small-bowel lymphoma | 1 | 75 | 1F | Hypogammaglobulinemia | NA | B-cell function deficiency | <1 year |
| 5 | Aguilar FP | 1987 | Jejunal lymphoma | 1 | 55 | 1F | Common variable hypogammaglobulinemia | NA | B-cell function deficiency | NA |
| 6 | Cunningham-Rundles C | 1987 | Gastric adenocarcinoma | 1 | 48 | 1F | CVID | NA | B-cell function deficiency | >5 years* |
| 7 | Durham JC | 1987 | Jejunal lymphoma | 1 | 52 | 1M | Common variable hypogammaglobulinemia | NA | B-cell function deficiency | <3 years |
| 8 | Castellano G | 1992 | Jejunal lymphoma | 1 | 39 | 1M | CVID | NA | B-cell function deficiency | <1 year |
| 9 | Chiaramonte C | 1994 | Jejunal lymphoma | 1 | 32 | 1F | CVID | NA | B-cell function deficiency | NA |
| 10 | Gabizon I | 2006 | Small-bowel carcinoid | 1 | 55 | 1F | Familial Mediterranean fever | *MEFV* | Autoinflammatory disorder | >5 years* |
| 11 | Pulvirenti F | 2018 | Small-bowel lymphoma | 3 | 49.6 | 1F, 2M | CVID | NA | B-cell function deficiency | Pt 1,2>5 years*  Pt 3>5 years |

*

Patients still alive at the time of publication were considered to have a survival of more than 5 years

**Supplementary Table 5. Patients with colorectal cancers**

| Paper  # | Reference first author | Year | Type of cancer | Case number | Age at cancer diagnosis | Sex | IEI type | Gene defect | Immune deficiency type | Prognosis |
| --- | --- | --- | --- | --- | --- | --- | --- | --- | --- | --- |
| 1 | Hermans PE | 1966 | Colon adenocarcinoma | 1 | 47 | 1F | CVID | NA | B-cell function deficiency | NA |
| 2 | Miller DG | 1967 | Colon adenocarcinoma | 1 | 46 | 1F | IgA deficiency | NA | B-cell function deficiency | NA |
| 3 | Kersey JH | 1973 | Colon adenocarcinoma | 1 | 12 | 1F | IgA deficiency | NA | B-cell function deficiency | NA |
| 4 | Hamoudi AB | 1974 | Colon adenocarcinoma | 1 | 20 | 1F | IgA deficiency | NA | B-cell function deficiency | <1 year |
| 5 | Hermans PE | 1976 | 1 Colon adenocarcinoma, 1 lymphoma | 2 | 57.5 | 1F, 1M | CVID | NA | B-cell function deficiency | Pt 1<1 year, Pt 2>5 years |
| 6 | Mir-Madjlessi SH | 1984 | Colon adenocarcinoma and lymphoma | 1 | 14 | 1M | IgA deficiency | NA | B-cell function deficiency | <1 year |
| 7 | Kinlen LJ | 1985 | Colon adenocarcinoma | 1 | 16 | 1NA | CVID | NA | B-cell function deficiency | 1NA |
| 8 | Cunningham-Rundles C | 1987 | Colon adenocarcinoma | 1 | 52 | 1M | CVID | NA | B-cell function deficiency | >5 years |
| 9 | Takemiya M | 1987 | Colon adenocarcinoma | 1 | 38 | 1M | Bloom syndrome | NA | Combined immunodeficiency | <1 year |
| 10 | Adachi Y | 1992 | Sigmoid and rectal adenocarcinoma | 1 | 22 | 1M | Other predominantly antibody deficiencies | NA | B-cell function deficiency | >5 years* |
| 11 | de Bruin NC | 1993 | Colon neuroendocrine carcinoma | 1 | 16 | 1M | CVID | NA | B-cell function deficiency | <1 year |
| 12 | Ohnoshi T | 1993 | Rectal non-Hodgkin lymphoma | 1 | 22 | 1M | CVID | NA | B-cell function deficiency | >5 years* |
| 13 | Van der Meer JW | 1993 | Colon adenocarcinoma | 3 | 30.7 | 3M | XLA | NA | B-cell function deficiency | Pt1,2 <1 year  Pt 3, NA |
| 14 | Zirkin HJ | 1996 | Colon small-cell undifferentiated carcinoma | 1 | 23 | 1M | Other predominantly antibody deficiencies | NA | B-cell function deficiency | <3 years |
| 15 | Murphy RC | 1999 | Colon adenocarcinoma | 1 | 15 | 1NA | Ataxia telangiectasia | NA | Combined immunodeficiency | <1 year |
| 16 | Wang J | 1999 | Colon adenocarcinoma | 1 | 37 | 1M | Bloom syndrome | NA | Combined immunodeficiency | <1 year |
| 17 | Balci S | 1999 | Colon adenocarcinoma | 1 | 17 | 1M | Bloom syndrome | *BLM* | Combined immunodeficiency | <3 years |
| 18 | Meyer S | 2004 | Perianal rhabdomyosarcoma | 1 | 7 | 1F | Nijmegen breakage syndrome | *NBS1* | Combined immunodeficiency | >5 years* |
| 19 | Purim O | 2007 | Colon adenocarcinoma | 1 | 56 | 1M | Familial Mediterranean fever | NA | Autoinflammatory disorder | >5 years* |
| 20 | Dunnigan M | 2007 | Colon lymphoma | 1 | 44 | 1M | CVID | NA | B-cell function deficiency | >5 years* |
| 21 | Brosens LAA | 2008 | Colon adenocarcinoma | 2 | 41 | 2M | XLA | *BTK* | B-cell function deficiency | NA |
| 22 | Thomas ERA | 2008 | Colon adenocarcinoma | 1 | 41 | 1F | Bloom syndrome | NA | Combined immunodeficiency | >5 years* |
| 23 | Benjazia E | 2011 | Colon adenocarcinoma | 1 | 19 | 1F | Bloom syndrome | NA | Combined immunodeficiency | <1year |
| 24 | Watkins C | 2012 | Rectal adenocarcinoma | 2 | 68 | 1F,1M | CVID | NA | B-cell function deficiency | Pt1,2<1 year |
| 25 | Goldberg A | 2016 | Rectal squamous cell carcinoma | 1 | 48 | 1M | CVID | NA | B-cell function deficiency | >5 years* |
| 26 | Schober T | 2017 | Colon smooth-muscle tumor | 2 | 5.5 | 2NA | CARMIL2 deficiency | *CARMIL2* | T-cell function deficiency | Pt1<1 year  Pt2 >5 years* |
| 27 | Pulvirenti F | 2018 | Colon lymphoma | 1 | 74 | 1F | CVID | NA | B-cell function deficiency | >5 years* |
| 28 | Antunes PDSL | 2019 | Colon adenocarcinoma | 1 | 10 | 1M | CVID | NA | B-cell function deficiency | <1 year |
| 29 | Li M | 2019 | Colon adenocarcinoma | 1 | 21 | 1M | XLA | *BTK* | B-cell function deficiency | <1 year |

*

Patients still alive at the time of publication were considered to have a survival of more than 5 years
